## Supplementary material for "Can the application of machine learning to electronic health records guide antibiotic prescribing decisions for suspected urinary tract infection in the Emergency Department?"

Affiliations:

### Systemic antibiotics recommended for UTI

The 2018 prescribing guidelines at Queen Elizabeth Hospital Birmingham recommended the following systemic antibiotics for the treatment of UTI in non-pregnant patients:

- **Lower UTI**
  - First line: nitrofurantoin (oral)
  - Second line: trimethoprim (oral)
- **Pyelonephritis**
  - First line: amoxicillin (iv), co-amoxiclav (oral), gentamicin (iv)
  - Second line: ciprofloxacin (oral), vancomycin (if history of UTI or suspected MRSA; iv), ertapenem (if history of UTI or suspected ESBL; iv)
- **(Uro)sepsis**
  - First line: co-amoxiclav (iv), gentamicin (iv)
  - Second line: ceftriaxone (iv), ciprofloxacin (iv), vancomycin (iv), piperacillin-tazobactam (iv), meropenem (if suspected ESBL or septic shock; iv)

Note that some of the above antibiotics are indicated in combination (e.g., co-amoxiclav and gentamicin as first line treatment for urosepsis). This was not enforced in this study and any prescription of one or more of the above antibiotics was considered as potential treatment for UTI for the purposes of this study.

### Missing data

EHR data is primarily recorded with patient management in mind. Information is only recorded if measuring *and* recording the information in the EHR system was prescribed by clinical guidelines or deemed necessary by the healthcare personnel in charge of patient care. Missing data is therefore ubiquitous when using EHR data for research. Negative information — i.e., information on the absence of an event — is seldom recorded explicitly, making it difficult to distinguish failure to record a disease from a genuine absence of disease. For example, a record of renal disease in a patient's medical history explicitly implies that a diagnosis has previously been made. Absence of renal disease, on the other hand, is usually implicitly signalled by an *absence* of diagnosis codes. As a consequence, we were unable to distinguish between cases in which a patient truly did not have a disease, and cases where the patient did suffer from the disease but the disease was not recorded, either because it hasn't been diagnosed yet or because the diagnosis was not captured in EHR records. For variables that describe the presence or absence of an event — i.e., those related to comorbidity diagnoses and past healthcare activity — we therefore assumed that an absence of a record meant that the event did not take place.

Missingness could be ascertained, however, in clinical information relating to general patient characteristics and clinical observations — i.e., demographic information, urine flow cytometry results, vital signs, and blood tests. For example, a missing record of heart rate can be unambiguously interpreted. We imputed likely values of these variables using several common imputation methods. These methods differed in their computational complexity but also statistical capability of faithfully reflecting the uncertainty caused by missing data. They may therefore differ in their impact on model performance. The following three imputation methods were applied to assess their impact on model performance:

**Mean imputation and missing indicators:** For mean imputation, each missing numerical value was replaced by the mean observed value in the training set. For categorical variables, missing values were assigned to a "missing" category. To allow the models to distinguish between patients with observed values and patients for whom the value was imputed with the mean, the models were additionally supplied with dummy variables that indicated the presence (0) or absence (1) of the value in the original, unimputed data.

**k-nearest neighbours imputation**: For k-nearest neighbours (KNN) imputation, each patient was matched with k patients most similar to him or her. Similarity between observations was estimated using Gower's distance, which is able to handle both continuous and categorical predictors (Gower 1971). Once the k neighbours were identified for a patient, his or her missing numerical values were imputed with the mean value among the k neighbours and categorical values were imputed with the mode. A value of k=5 was chosen for this analysis.

**Multiple imputation using multivariate imputation by chained equations**: All previous imputation methods produce only a single imputation for each missing value. The prediction model would treat those values as if they were actually observed, ignoring the inherent uncertainty involved in the imputation process (White et al. 2011). Multiple imputation addresses this issue and imputes each missing value with M random draws from an imputation model — e.g., linear regression or predictive mean matching — resulting in M imputed datasets. A separate prediction model was then fit to each imputed dataset, and the M predictions averaged across datasets. By considering multiple plausible imputations, multiple imputation actively accounts for the uncertainty surrounding the imputation process. Imputation was performed using the multivariate imputation by chained equations (MICE) algorithm with predictive mean matching (continuous variables), logistic regression (binary variables), and multinomial regression (categorical variables) (White et al. 2011; van Buuren 2018). Five datasets were imputed, and the algorithm was run for 10 iterations to achieve approximate convergence. All variables were assumed to be missing at random, meaning that the probability of a value being missing depended only on the values of observed covariates. An example of this missingness mechanism would be if creatinine were less frequently measured in younger, otherwise healthy patients, and the probability of it being measured only depended on age and comorbidities (which are covariates in the data). Due to computational restrictions in the safe haven in which the analysis was performed, multiple imputation was only performed for logistic regression models.

Imputation was performed separately in each resampled dataset (i.e., in each iteration of each [repeated] 10-fold cross validation). Each imputation strategy was trained on the split's training data (i.e., the combined 9 training folds of the iteration) and applied without retraining to the split's test data (i.e., the held-out test fold of the iteration). For multiple imputation, we included the outcome as a predictor in the imputation model as recommended (Moons et al. 2006). Since the outcome won't be available during model deployment — i.e., in real-time on the hospital ward — a second set of multiple imputed datasets was imputed *without* the outcome which was then used on the held-out test fold. This ensured that the evaluation faithfully reflected the eventual intended use of the model (Wood et al. 2015; Rockenschaub et al. 2020).

### Testing differences in model performance

Model performance may depend on the exact case mix in the training and test sets. We therefore evaluated model performance using resampling techniques in both internal validation (repeated cross-validation) and external validation (bootstrap).

Cross-validation was chosen over bootstrap during internal validation, since the (optimism-adjusted) bootstrap originally proposed in (Rockenschaub et al. 2020) was found inadequate to prevent overfitting for very flexible models such as random forests and gradient boosting trees. To account for the dependence introduced within a single cross-validation split (i.e., a sample that is in fold 1 cannot be in fold 2), differences in model performances were tested for statistical significance using Bayesian generalised linear mixed models estimated via Markov Chain Monte Carlo sampling with four chains of 2,000 warm-up iterations and 2,000 sampling iterations (Benavoli et al. 2017).

Since external validation did not require additional model fitting on the test data (the model is already fit on the entire training data), we chose simple bootstrapping to obtain an estimate of the variability of our performance estimates in the test data. These bootstraps do not suffer from the same dependence that might arise from repeated cross-validation. Confidence intervals were therefore estimated directly from 1,000 bootstraps as the proportion of bootstraps in which model A achieved higher performance than model B, multiplied by two to account for the two-sided nature of our hypothesis.

### Supplementary Tables

Supplementary Table 1. Discriminative performance of candidate models when predicting bacterial growth during 10-times repeated 10-fold cross-validation of the development set.

| **Model** | **AUC (95% CI)** | **Specificity (95% CI)** | **NPV (95% CI)** | **p-value** |
|---|---|---|---|---|
| All candidate predictors | | | | |
| XGB | 0.808 (0.805-0.811) | 34.9 (33.9-35.9) | 92.4 (92.2-92.7) | - |
| RF | 0.804 (0.802-0.807) | 35.4 (34.5-36.4) | 92.5 (92.3-92.7) | <0.001 |
| E-NET | 0.788 (0.785-0.791) | 29.7 (28.8-30.5) | 91.2 (91.0-91.5) | <0.001 |
| LR | 0.785 (0.782-0.788) | 28.7 (27.8-29.5) | 90.9 (90.7-91.2) | <0.001 |
| LR-FP | 0.781 (0.778-0.784) | 28.3 (27.4-29.2) | 90.8 (90.5-91.1)) | <0.001 |
| Reduced set of predictors | | | | |
| XGB | 0.795 (0.792-0.798) | 34.3 (33.5-35.2) | 92.4 (92.2-92.6) | <0.001 |
| E-NET | 0.773 (0.770-0.777) | 28.6 (27.9-29.4) | 90.8 (90.6-91.0) | <0.001 |
| LR | 0.773 (0.769-0.776) | 28.8 (28.1-29.5) | 90.9 (90.6-91.0) | <0.001 |
| LR-FP | 0.768 (0.765-0.771) | 27.5 (26.7-28.2) | 90.4 (90.2-90.7) | <0.001 |
| RF | 0.767 (0.764-0.770) | 12.2 (11.7-12.6) | 77.5 (76.7-78.4)) | <0.001 |

Specificity and NPV were calculated at a predefined sensitivity of 95%. p-values were obtained via Bayesian generalised linear mixed models (Benavoli et al. 2017).

AUC, area under the receiver operating characteristic; CI, confidence interval; E-NET, elastic net; LR, logistic regression; LR-FP, logistic regression with fractional polynomials; NPV, negative predictive value; RF, random forest; XGB, extreme gradient boosting trees.

Supplementary Table 2. Estimated AUC of LR and XGB using all predictors during external validation, by imputation method.

| **Imputation method** | **Model** | |
|---|---|---|
| | **LR** | **XGB** |
| | AUC (95% CI) | AUC (95% CI) |
| Mean | 0.796 (0.776-0.817) | 0.813 (0.792-0.834) |
| kNN | 0.796 (0.776-0.817) | 0.814 (0.794-0.835) |
| MICE (M=5) | 0.769 (0.742-0.796) | * |

* Not calculated due to computational limitations in the data safe haven within which the analysis was performed.

AUC, area under the receiver operating characteristic; CI, confidence interval; kNN, k-nearest neighbours; LR, logistic regression; MICE; multivariate imputation by chained equations; XGB, extreme gradient boosting trees.

Supplementary Table 3. Top ten variables with the highest AUC when predicting bacterial growth during internal validation of LR and XGB.

| **LR** | | | | **XGB** | | |
|---|---|---|---|---|---|---|
| **#** | **Variable** | **AUC (95% CI)** | | **#** | **Variable** | **AUC (95% CI)** |
| 1 | UFC bacteria | 0.631 (0.625-0.636) | | 1 | UFC bacteria | 0.662 (0.657-0.667) |
| 2 | ED diagnosis | 0.603 (0.598-0.608) | | 2 | UFC white blood cells | 0.608 (0.602-0.614) |
| 3 | UFC epithelial cells | 0.585 (0.577-0.593) | | 3 | ED diagnosis | 0.603 (0.598-0.609) |
| 4 | UFC white blood cells | 0.581 (0.575-0.587) | | 4 | UFC epithelial cells | 0.580 (0.573-0.588) |
| 5 | UFC casts | 0.561 (0.556-0.566) | | 5 | UFC casts | 0.560 (0.555-0.565) |
| 6 | UFC small round cells | 0.555 (0.549-0.562) | | 6 | UFC small round cells | 0.559 (0.552-0.565) |
| 7 | UFC red blood cells | 0.549 (0.543-0.556) | | 7 | Age | 0.548 (0.542-0.553) |
| 8 | Age | 0.548 (0.542-0.553) | | 8 | Missing UFC | 0.546 (0.539-0.554) |
| 9 | Missing UFC | 0.546 (0.539-0.554) | | 9 | UFC red blood cells | 0.543 (0.535-0.550) |
| 10 | Previous bacteriuria | 0.539 (0.534-0.545) | | 10 | Previous bacteriuria | 0.539 (0.534-0.545) |

AUC, area under the receiver operating characteristic; CI, confidence interval; ED, emergency department; LR, logistic regression; UFC, urine flow cytometry; XGB, extreme gradient boosting trees.

Supplementary Table 4. Comparison of demography, medical history, and clinical characteristics between training set (before or in 2017) and test set (after 2017).

| | Training set | | Test set | | p-value |
|---|---|---|---|---|---|
| | Summary | Missing % | Summary | Missing % | |
| Number of visits | 10,352 (100.0) | | 1,538 (100.0) | | |
| **Demographics** | | | | | |
| ≥65 years (%) | 6,142 (51.7) | 0.0 | 5,382 (52.0) | 0.0 | 0.063 |
| Female (%) | 7,851 (66.0) | 0.0 | 6,838 (66.1) | 0.0 | 0.906 |
| Ethnicity (%) | | | | | 0.035 |
| Asian | 1,342 (13.9) | | 221 (16.1) | | |
| Black | 447 (4.6) | | 68 (4.9) | | |
| White | 7,666 (77.1) | | 1,019 (74.1) | | |
| Other | 434 (4.5) | | 68 (4.9) | | |
| **Comorbidities** | | | | | |
| Charlson comorbidity index (%) | | 0.0 | | 0.0 | 0.016 |
| 0 | 6,089 (58.8) | | 956 (62.2) | | |
| 1-2 | 2,482 (24.0) | | 320 (20.8) | | |
| ≥3 | 1,781 (17.2) | | 262 (17.0) | | |
| Cancer (%) | 714 (6.9) | 0.0 | 126 (8.2) | 0.0 | 0.072 |
| Underlying renal condition (%) | 2,134 (20.6) | 0.0 | 304 (19.8) | 0.0 | 0.078 |
| Underlying urological condition (%) | 2,902 (28.0) | 0.0 | 336 (21.8) | 0.0 | 0.462 |
| Renal/urological surgery (%) | 1,985 (19.2) | 0.0 | 256 (16.6) | 0.0 | 0.020 |
| **Hospital activity in prior year** | | | | | |
| Any hospitalisation (%) | 4,910 (47.4) | 0.0 | 667 (44.0) | 0.0 | 0.013 |
| Urine sample taken (%) | 5,097 (49.2) | 0.0 | 623 (40.5) | 0.0 | <0.001 |
| Urine sample positive (%) | 2,476 (23.9) | 0.0 | 306 (19.9) | 0.0 | 0.001 |
| Antibiotics in hospital (%) | 2,497 (24.1) | 0.0 | 366 (23.8) | 0.0 | 0.806 |
| **Presentation in the ED** | | | | | |
| Recorded ED diagnosis (%) | | 4.1 | | 18.4 | <0.001 |
| UTI | 4,054 (39.2) | | 639 (41.5) | | |
| UTI symptoms | 1,554 (15.0) | | 112 (7.3) | | |
| Other infection | 1,339 (12.9) | | 228 (14.8) | | |
| Other diagnoses | 2,978 (28.8) | | 276 (17.9) | | |
| Urine flow cytometry (median / IQR) | | | | | |
| Bacteria x$10^3$/µL | 9.0 (3.0, 24.)] | 13.3 | 3.4 (0.5, 11.1) | 24.9 | <0.001 |
| White blood cells x1/µL | 311 (103, 1154) | 13.3 | 410 (157, 1321) | 24.9 | <0.001 |
| Red blood cells x1/µL | 38.0 (14.0, 151.0) | 13.3 | 24.0 (10.0, 76.5) | 24.9 | <0.001 |
| Epithelial cells x1/µL | 23.0 (8.0, 58.5) | 13.3 | 11.0 (3.0, 32.5) | 24.9 | <0.001 |
| Small round cells x1/µL | 2.0 (1.0, 4.0) | 13.2 | 1.0 (0.0, 3.0) | 24.9 | <0.001 |
| Casts x1/µL | 1.0 (0.0, 2.0) | 13.2 | 1.0 (0.0, 2.0) | 24.9 | 0.021 |
| Crystals x1/µL | 5.0 (2.0, 15.0) | 51.1 | - | 100.0 | |
| Blood tests (median / IQR) | | | | | |
| C-reactive protein mg/L | 27.0 (6.0, 94.0) | 56.4 | 34.0 (8.0, 94.0) | 43.5 | 0.050 |
| White blood cells x$10^3$/µL | 10.8 (8.1, 14.5) | 46.3 | 10.8 (8.0, 14.5) | 34.3 | 0.907 |
| Platelets x$10^3$/µL | 229 (181, 290) | 46.4 | 238 (190, 302) | 34.5 | <0.001 |
| Creatinine µmol/L | 84.0 (66.0, 119) | 48.0 | 79.5 (64.0, 109) | 33.6 | <0.001 |
| Bilirubin µmol/L | 9.0 (6.0, 14.0) | 54.7 | 9.0 (6.0, 14.0) | 36.5 | 0.989 |
| Alkaline phosphatase IU/L | 87.0 (68.0, 117) | 54.3 | 88.0 (69.0, 117) | 38.3 | 0.789 |
| **Outcome** | | | | | |
| Bacterial growth observed | 3,850 (35.5) | 0.0 | 678 (44.1) | 0.0 | <0.001 |

ED, emergency department; IQR, interquartile range.

Supplementary Table 5. Applied Yeo-Johnson transformations and final model coefficients for baseline LR models using all predictors or the reduced set of predictors.

| | **Yeo-Johnson transformation** | | | **LR All predictors** | **LR Reduced predictors** |
|---|---|---|---|---|---|
| | Lambda | Mean | SD | OR (95% CI) | OR (95% CI) |
| Base probability of **no** growth* | | | | 87.5% (72.5-94.9%)* | 77.4% (74.1-80.5%)* |
| **Demographics** | | | | | |
| Age in years | | | | | |
| 18 - 25 | | | | 1 | 1 |
| 25 - 34 | | | | 1.15 (0.95-1.39) | 1.26 (1.05-1.52) |
| 35 - 44 | | | | 0.98 (0.79-1.21) | 1.15 (0.93-1.41) |
| 45 - 54 | | | | 0.82 (0.66-1.02) | 1.06 (0.86-1.30) |
| 55 - 64 | | | | 0.95 (0.77-1.19) | 1.21 (0.98-1.49) |
| 65 - 74 | | | | 0.81 (0.66-1.00) | 1.07 (0.89-1.29) |
| 75 - 84 | | | | 0.95 (0.78-1.15) | 1.29 (1.08-1.54) |
| 85 - 94 | | | | 0.90 (0.74-1.10) | 1.26 (1.05-1.50) |
| 95 - 104 | | | | 1.18 (0.81-1.72) | 1.64 (1.14-2.35) |
| Female | | | | 0.64 (0.57-0.73) | 0.57 (0.51-0.63) |
| Ethnicity | | | | | |
| White | | | | 1 | |
| Asian | | | | 1.04 (0.90-1.21) | |
| Black | | | | 1.08 (0.86-1.37) | |
| Other | | | | 1.00 (0.78-1.27) | |
| Unknown | | | | 0.94 (0.75-1.18) | |
| **Comorbidities** | | | | | |
| Charlson comorbidity index (linear) | | | | 1.01 (0.98-1.05) | |
| Cancer | | | | 0.88 (0.70-1.11) | |
| Underlying renal condition | | | | 0.90 (0.78-1.05) | |
| Underlying urological condition | | | | 1.04 (0.90-1.21) | |
| Renal/urological surgery | | | | 1.24 (1.07-1.44) | |
| **Hospital activity** | | | | | |
| Hospital activity in prior 7 days | | | | 0.93 (0.74-1.16) | |
| Number of hosp. in prior year | | | | 0.98 (0.94-1.02) | |
| Number of UTI hosp. in prior 2 years | | | | 1.06 (0.96-1.17) | |
| Number of ED visits in prior year | | | | 1.00 (0.97-1.03) | |
| Number of UTI ED visits in prior 2 years | | | | 1.11 (1.01-1.22) | |
| Urine culture in prior year | | | | 1.44 (1.26-1.64) | |
| Positive urine culture in prior year | | | | 0.62 (0.54-0.71) | 0.87 (0.78-0.96) |
| Antibiotics in prior year | | | | 1.08 (0.92-1.26) | |
| **Presentation in the ED** | | | | | |
| Recorded ED diagnosis (%) | | | | | |
| UTI | | | | 1 | |
| Pyelonephritis | | | | 0.77 (0.62-0.95) | |
| Urosepsis | | | | 1.57 (1.26-1.95) | |
| Urinary symptoms | | | | 1.90 (1.49-2.42) | |
| Altered mental status | | | | 1.71 (1.35-2.17) | |
| Abdominal pain | | | | 2.43 (1.87-3.16) | |
| Other sepsis | | | | 2.40 (1.82-3.17) | |
| Lower resp. tract infection | | | | 2.23 (1.73-2.87) | |
| Other infections | | | | 2.04 (1.60-2.62) | |
| Other genitourinary conditions | | | | 1.72 (1.24-2.40) | |
| Other diagnoses | | | | 1.71 (1.51-1.95) | |
| Urine flow cytometry (median / IQR) | | | | | |
| Bacteria x$10^3$/μL | 0.0590 | 11.80 | 3.137 | 0.34 (0.32-0.37) | 0.35 (0.33-0.37) |

| | | | | | |
|---|---|---|---|---|---|
| White blood cells x1/μL | -0.0357 | 5.25 | 1.286 | 0.63 (0.60-0.67) | 0.61 (0.58-0.64) |
| Red blood cells x1/μL | -0.2183 | 2.55 | 0.644 | 1.19 (1.13-1.26) | 1.20 (1.13-1.26) |
| Epithelial cells x1/μL | -0.0078 | 3.08 | 1.255 | 1.72 (1.60-1.85) | 1.72 (1.60-1.85) |
| Small round cells x1/μL | -0.3953 | 0.80 | 0.540 | 0.87 (0.82-0.92) | 0.87 (0.82-0.93) |
| Casts x1/μL | -0.6553 | 0.49 | 0.418 | 1.22 (1.14-1.31) | 1.25 (1.16-1.33) |
| Crystals x1/μL | -0.2442 | 1.46 | 0.564 | 1.07 (1.02-1.12) | 1.07 (1.02-1.12) |
| Blood tests (median / IQR) | | | | | |
| C-reactive protein mg/L | -0.0497 | 3.01 | 0.806 | 0.96 (0.91-1.01) | |
| White blood cells x10$^3$/μL | 0.0149 | 2.52 | 0.311 | 0.92 (0.87-0.97) | |
| Platelets x10$^3$/μL | 0.4399 | 22.7 | 3.122 | 1.10 (1.05-1.17) | |
| Creatinine μmol/L | -0.7859 | 1.23 | 0.009 | 1.04 (0.98-1.09) | |
| Bilirubin μmol/L | -0.4495 | 1.42 | 0.138 | 1.07 (1.01-1.13) | |
| Alkaline phosphatase IU/L | -0.7054 | 1.36 | 0.012 | 1.00 (0.95-1.05) | |
| Missing values | | | | | |
| All flow cytometry missing | | | | 0.98 (0.83-1.16) | 1.02 (0.87-1.21) |
| Some flow cytometry missing | | | | 0.44 (0.38-0.51) | 0.46 (0.39-0.53) |
| Only casts missing | | | | 1.17 (0.98-1.41) | 1.17 (0.98-1.40) |
| WBC (blood) missing | | | | 0.92 (0.77-1.10) | |
| Creatinine count missing | | | | 0.95 (0.70-1.28) | |
| Alk. phosph. and bili. missing | | | | 1.03 (0.81-1.30) | |
| C-reactive protein missing | | | | 1.04 (0.87-1.25) | |
| **Time of arrival** | | | | | |
| Month | | | | | |
| January | | | | 1 | |
| February | | | | 0.93 (0.70-1.25) | |
| March | | | | 0.65 (0.44-0.96) | |
| April | | | | 0.55 (0.32-0.94) | |
| May | | | | 0.48 (0.24-0.94) | |
| June | | | | 0.31 (0.14-0.72) | |
| July | | | | 0.36 (0.14-0.98) | |
| August | | | | 0.26 (0.08-0.83) | |
| September | | | | 0.20 (0.05-0.74) | |
| October | | | | 0.16 (0.04-0.70) | |
| November | | | | 0.13 (0.02-0.64) | |
| December | | | | 0.12 (0.02-0.73) | |
| Day of the year (linear) | | | | 1.01 (1.00-1.01) | |
| Day of the week | | | | | |
| Monday | | | | 1 | |
| Tuesday | | | | 1.10 (0.93-1.30) | |
| Wednesday | | | | 1.08 (0.91-1.28) | |
| Thursday | | | | 1.02 (0.86-1.21) | |
| Friday | | | | 1.17 (0.98-1.38) | |
| Saturday | | | | 1.24 (1.04-1.49) | |
| Sunday | | | | 1.18 (1.00-1.40) | |
| Time of day (linear) | | | | 1.00 (0.99-1.01) | |

* Note that the tidymodels framework per default sets the positive event (=culture growth) as the reference category. This differs from most other statistical packages, and the LR model therefore estimates the probability of *no* growth for a reference patient. To give a concrete example, the reduced model estimates that 77.4% of reference patients will show *no* growth and that women are 0.57 as likely to show no growth (i.e., they are more likely to show growth). The probability of growth can be obtained via 100-77.4=22.6% and odds ratio for growth in females compared to males can be obtained via 1 / 0.57 = 1.75. Reference patients have mean valuesfor each continuous variable and each categorical variable set to the reference category. For example, for the reduced model this is a 18 - 25 year old male with no positive urine sample in the prior 12 months, no missing urine flow cytometry data, and average values for the flow cytometry measurements.

### Supplementary Figures

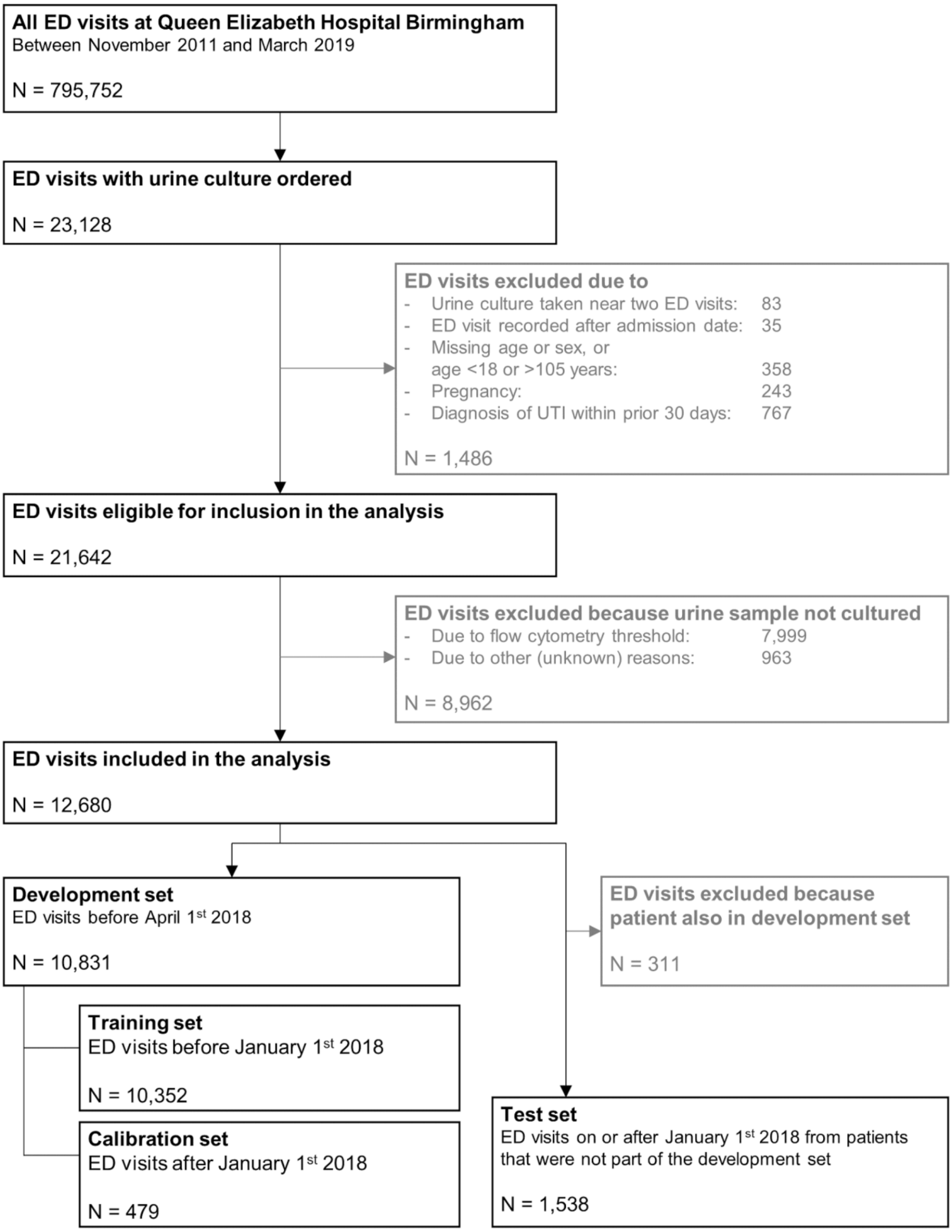


Supplementary Figure 1 - Flow chart of cohort selection for community-onset UTI in the ED at QEHB.

ED, emergency department; QEHB, Queen Elizabeth Hospital Birmingham; UTI, urinary tract infection.

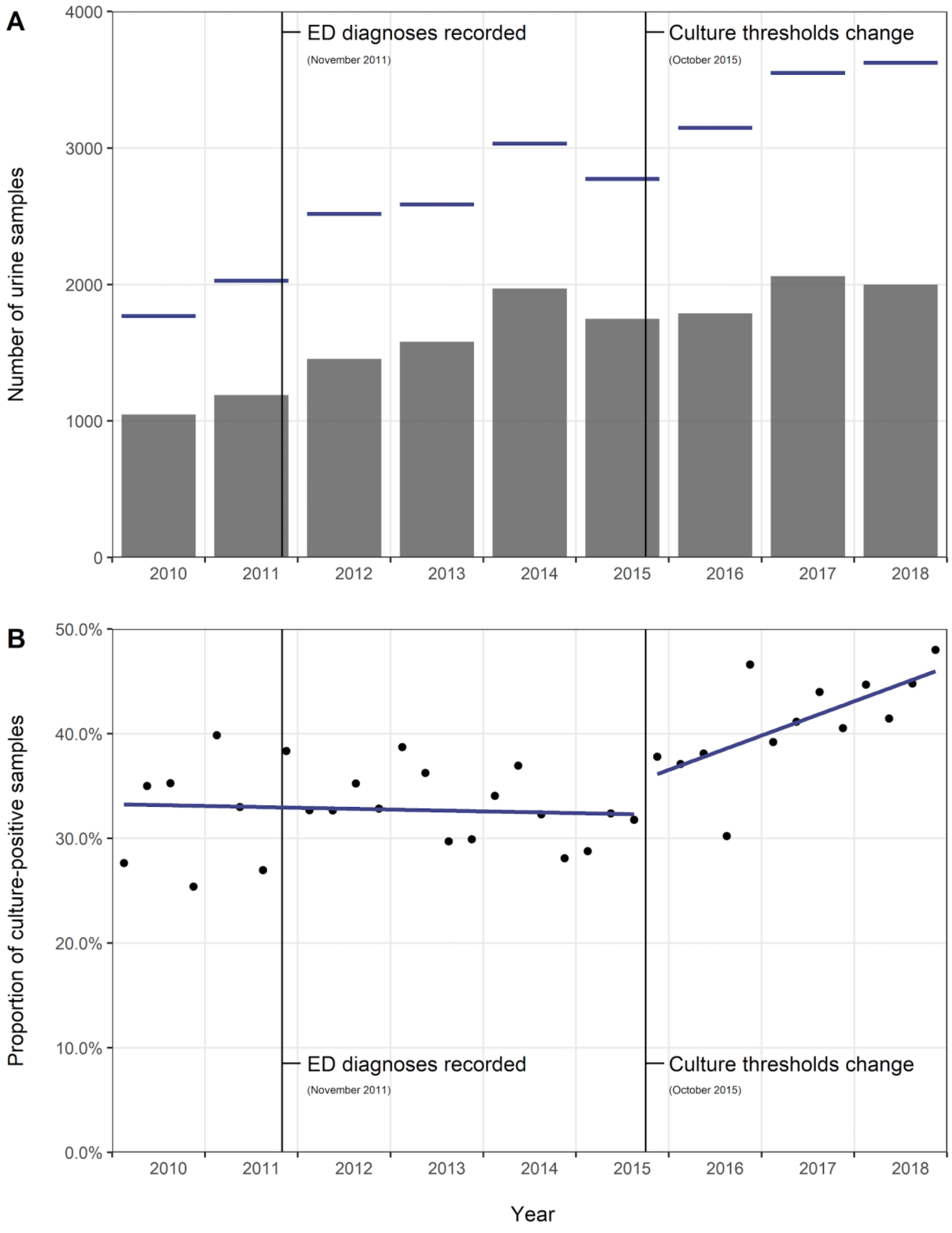


Supplementary Figure 2 - A) Yearly distribution of ED visits with a urine sample sent for microbiological culture (blue lines), and number of ED visits for which the urine sample was ultimately cultured (grey bars). Although visits before November 2011 are presented here to show an overall trend, they were not included in the main analysis since ED diagnoses were not yet recorded for these visits. B) Quarterly proportion of cultured urine samples that showed predominant bacterial growth (black dots) and linear trend (blue lines) before and after the change in urinalysis thresholds in October 2015.

ED, emergency department.

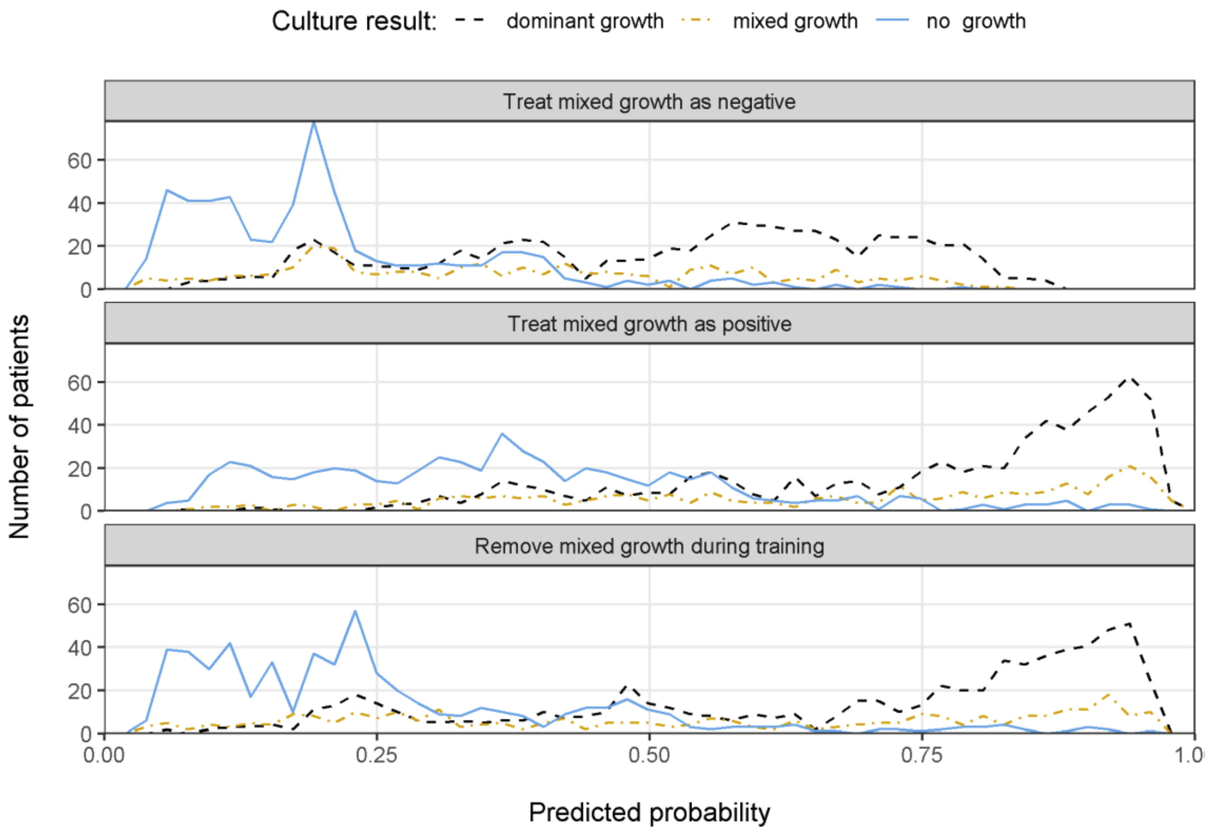


Supplementary Figure 3 - Distribution of model predictions in the test set for samples with dominant growth, mixed growth, and no growth, depending on how mixed growth was treated during model training.
