## Supplementary material for "Can the application of machine learning to electronic health records guide antibiotic prescribing decisions for suspected urinary tract infection in the Emergency Department?": Confirmation of waived ethics

Go straight to content.

Medical Research Council

NHS Health Research Authority

### Do I need NHS REC review?

**To print your result with title and IRAS Project ID please enter your details below:**

Title of your research:

Precision prescribing for UTI in hospital

IRAS Project ID (if available):

Your answers to the following questions indicate that **you do not need NHS REC review for sites in England.**

This tool only considers whether NHS REC review is required, it does not consider whether other approvals are needed. You should check what other approvals are required for your research.

You have answered **'YES'** to: Is your study research?

You answered **'NO'** to all of these questions:

**Question Set 1**

- Is your study a clinical trial of an investigational medicinal product?
- Is your study one or more of the following: A non-CE marked medical device, or a device which has been modified or is being used outside of its CE mark intended purpose, and the study is conducted by or with the support of the manufacturer or another commercial company (including university spin-out company) to provide data for CE marking purposes?
- Does your study involve exposure to any ionising radiation?
- Does your study involve the processing of disclosable protected information on the Register of the Human Fertilisation and Embryology Authority by researchers, without consent?

**Question Set 2**

- Will your study involve potential research participants identified in the context of, or in connection with, their past or present use of services (NHS and adult social care), including participants recruited through these services as healthy controls?
- Will your research involve prospective collection of tissue (i.e. any material consisting of or including human cells)

from any past or present users of these services (NHS and adult social care)?
- Will your research involve prospective collection of information from any past or present users of these services (NHS and adult social care)?
- Will your research involve the use of previously collected tissue and/or information from which individual past or present users of these services (NHS and adult social care), are likely to be identified by the researchers either directly from that tissue or information, or from its combination with other tissue or information likely to come into their possession?
- Will your research involve potential research participants identified because of their status as relatives or carers of past or present users of these services (NHS and adult social care)?

**Question Set 3**

- Will your research involve the storage of relevant material from the living or the deceased on premises in England, Wales or Northern Ireland without a storage licence from the Human Tissue Authority (HTA)?
- Will your research involve storage or use of relevant material from the living, collected on or after 1st September 2006, and the research is not within the terms of consent for research from the donors?
- Will your research involve the analysis of human DNA in cellular material (relevant material), collected on or after 1st September 2006, and this analysis is not within the terms of consent for research from the donor? And/or: Will your research involve the analysis of human DNA from materials that do not contain cells (for example: serum or processed bodily fluids such as plasma and semen) and this analysis is not within the terms of consent for research from the donor?

**Question Set 4**

- Will your research involve at any stage procedures (including use of identifiable tissue samples or personal information) involving adults who lack capacity to consent for themselves, including participants retained in study following the loss of capacity?
- Is your research health-related and involving offenders?
- Does your research involve xenotransplantation?
- Is your research a social care project funded by the Department of Health and Social Care (England)?
- Will the research involve processing confidential information of patients or service users outside of the care team without consent? And/ or: Does your research have Section 251 Support or will you be making an application to the Confidentiality Advisory Committee (CAG) for Section 251 Support?

If your research extends beyond **England** find out if you need NHS REC review by selecting the 'OTHER UK COUNTRIES' button below.

OTHER UK COUNTRIES

**If, after visiting all relevant UK countries, this decision tool suggests that you do not require NHS REC review follow this link for final**

**confirmation and further information.**

Print This Page

NOTE: If using Internet Explorer please use browser print function.

About this tool Feedback Contact Glossary Algorithm
Accessibility
